# Perceived barriers to research and scholarship among physicians

**DOI:** 10.1101/2024.09.16.24313581

**Authors:** Makayla Lagerman, Lauren Dennis, Traci Deaner, Adam Sigal, David Rabago, Huamei Dong, Joseph P. Wiedemer, Alexis Reedy-Cooper, Jessica Parascando, Karl T. Clebak, Tara Cassidy-Smith, Robert P. Lennon, Olapeju Simoyan

## Abstract

**Introduction:** Inadequate scholarship is a common concern in graduate medical education. Many barriers to scholarship have been identified, but there are limited data on which are most important. A rank-order of barriers from learners’ perspectives would better enable programs to address perceived barriers.

**Methods:** Given that learners are experts in their own perceptions, we applied the Delphi method of generating consensus expert opinion to construct ranked lists of physician-perceived barriers to scholarly activity at various sites. The survey was conducted within three separate health systems. Respondents were asked to identify their perceived barriers via free text and the listed barriers were consolidated by the research team. In the second round, respondents were presented with the consolidated lists and asked to rank them. In the third and final round, each respondent was shown a comparison of their own rankings to that of their peers and given an opportunity to make changes. Ranking differences between programs were compared using Rank Biased Overlap (RBO).

**Results:** The Delphi method is a straightforward method to obtain a ranked list of perceived learner barriers to scholarly activity; its primary limitation is learner engagement and, of note in our study, high dropout rates. RBO is an effective method of ranking differences between programs and specialties. Top barriers across programs included time, overwhelm with the research process, and lack of interest or energy.

**Discussion:** Several of the identified barriers may be addressed with enthusiastic mentors, streamlined administrative processes, and education. This could be done within a hospital system or on a national level through specialty organizations.

## Introduction

Scholarly activity (SA) is a required component of graduate medical training and practice. For example, medical residents in family medicine are required to participate in and complete at least two scholarly activities before graduation.^1^ At academic medical centers, fellows and faculty are also expected to participate in research or scholarship.^2^ Barriers to resident scholarship have been identified^3,4^ but not ranked. Ranking of identified barriers may enable programs to prioritize interventions and determine if unique strategies in other programs are likely to work in their own.^1,5^ Barriers that are ranked highly by several programs may indicate a need for specialty-wide interventions through Accreditation Council for Graduate Medical Education (ACGME) program requirements or online resources.

The Delphi method is an iterative method of developing consensus among experts, often used in health and social sciences to identify intra-professional opinions.^6^ In medical education, the Delphi method has been used to obtain consensus opinion on content and methods of education and training.^7^ It has also been used to develop an electrocardiogram teaching curriculum for medical residents.^8^ It has not been used to identify a rank-order list of barriers to scholarly activity among medical learners. The Delphi technique allows participants to reflect and reconsider their opinions, based on the opinions of others.^9^ The Delphi method would be appropriate for this study since physicians are best able to identify their own perceived barriers to scholarship.

The utility of ranked lists of barriers to scholarly activity is increased if lists between programs can be compared. This may be challenging when using the Delphi method, as the identified barriers will differ in number and type between programs, precluding use of the Kendall Tau or Spearman’s Rank-Order Correlation methods. Rank Biased Overlap (RBO) compares lists of different elements by weighting each rank position.^10^ This has the added benefit of enabling greater weight to be placed on the top-ranked items of a given comparison. To our knowledge, RBO has not been applied to compare ranked scholarly activity barriers between programs.

The objective of this study was to identify perceived barriers to scholarly activity by conducting three-round Delphi surveys of medical residents, fellows, and faculty across several sites. The RBO method was used to compare ranked barriers between programs and determine if perceived barriers to scholarship differed by specialty or health system.

## Methods

Approval for this study was obtained from the System 1 (S1), System 2 (S2), and System 3 (S3) Institutional Review Boards.

### Delphi method

In the first survey, participants were asked to identify barriers to their scholarly activity via free text. Perceived barriers were consolidated; for example, “Not enough time” and “Too little time” were combined into “Time”. In the second round, participants were asked to rank the consolidated barriers. In the third round, participants were shown their rankings and the peer-average ranking for each barrier and allowed to modify their rankings. The consolidations occurred based on site and/or specialty.

### System 1

Site 1 is an academic medical system that includes a 714-bed hospital in an urban setting and 230 external locations, through which it provides primary and specialty care and graduate medical education to over 200 residents and fellows across several specialties. We distributed a three-round Delphi survey electronically to medical residents, fellows, and faculty at S1 from February through May 2021.

### System 2 Family Medicine residency programs

S2 has three family medicine residency programs. The S2-A program (8/8/8) is embedded in a 628-bed academic facility with dozens of specialty residencies. S2-B program (6/6/6) is unopposed and embedded in a 260-bed community medical center. S2-C program (6/6/6) is also unopposed and embedded in a 204-bed acute care hospital.

We distributed a three-round Delphi survey electronically to residents between August and September 2021 at three S2 Family and Community Medicine residencies. Each program completed their Delphi surveys independently. Program Directors were asked to provide dedicated time during weekly academic sessions to complete the survey.

### System 2-A and System 3 surgical and non-surgical residency programs

S2-A and S3 are both tertiary academic medical centers with 628 and 663 beds respectively. S2-A is located in central Pennsylvania, and S3 is located in urban New Jersey. Program Directors were asked to provide dedicated time during weekly academics to complete the survey.

We distributed an electronic three-round Delphi survey to residents at S2-A and S3between August and September 2021. Participants at all sites who completed all three surveys were eligible to enter a raffle to receive a $10 or $50 gift card.

### Rank Biased Overlap

Rank Biased Overlap (RBO) has an adjustable parameter to weight differences based on top order, so that lists with high overlap on highly ranked items (e.g., lists where the top five ranked items are the same) do not appear dissimilar because of less similarity on lower ranked items.^10^ As this is the first use of RBO to compare ranked lists using the Delphi method for this purpose, there is no defined parameter for optimal weighting. We therefore used a series of weighting parameters to explore the extent of similarity between lists.

## Results

### System 1

Of the 222 residents, fellows, and faculty invited to participate at S1, 72 completed the first survey (32% response rate). After the second survey was sent out, 31 respondents completed it (57% dropout rate). Nineteen respondents competed the final survey (73% dropout rate). Table 1 shows the top barriers, which include lack of understanding of process, tedious IRB process, and lack of time, from a mixed cohort of S1 residents, fellows, and faculty.

**Table 1.**
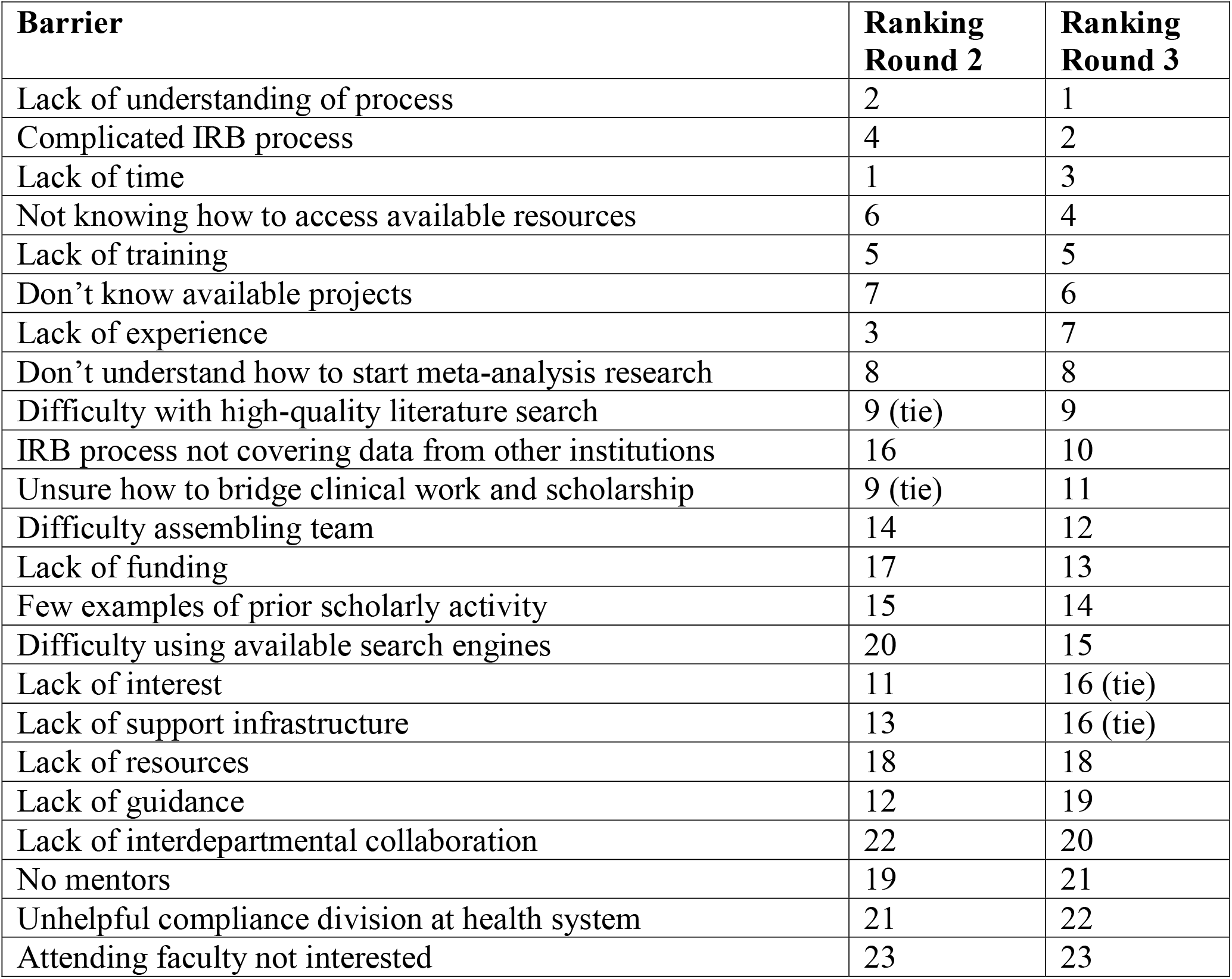
Ranked barriers to scholarly activity at System 1. Top ranked barriers after the third round were lack of understanding of process, complicated IRB process, and lack of time.

### System 2 Family Medicine residency programs

Of the 61 family medicine residents invited to participate, 29 completed the first survey (48% response rate). Response rates and barriers varied by site: S2-A 33%, 14 barriers; S2-B 42%, 15 barriers; S2-C 72%, 25 barriers. Only ten residents completed the second survey (66% dropout rate), and dropout rates varied by site: S2-A 38%; S2-B 50%; S2-C 92%. S2-C was not compared to the other sites as only one resident remained. Six residents completed the third survey; dropout rates were 63% for both S2-A and S2-B.

Table 2 shows the barriers and family medicine residents’ rankings for rounds 2 and 3. Top barriers include lack of time, lack of interest, and tedious research process. Table 3 shows the RBO for weighting parameters from 0.4 to 0.9 comparing S2-A and S2-B.

**Table 2.** System 2 Family Medicine residents’ ranked barriers to scholarly activity per site using Delphi method. Time was the top-ranked barrier at sites S2-A and S2-B. Site S2-C had only one respondent, preventing a third round rank.

| S2-A |  |  | S2-B |  |  | S2-C |  |
| --- | --- | --- | --- | --- | --- | --- | --- |
| Barriers | Ranking Round |  | Barriers | Ranking Round |  | Barriers | Ranking Round |
|  | 2 | 3 |  | 2 | 3 |  | 2 |
| Time | 1 | 1 | Time | 1 | 1 | No (personal) interest in research | 1 |
| Energy during residency when exhausted and busy | 4 | 2 | No interest in scholarship/lack of will | 2 | 2 | Bureaucracy / Paperwork is overwhelming | 2 |
| Prioritizing other learning experiences | 2 | 3 | Lack of training: IRB | 3 | 3 | Forced to be on projects not interested in | 3 |
| Overwhelming logistics, including IRB approval process, paperwork | 3 | 4 | Lack of knowledge: getting published | 4 | 4 | Lack of knowledge: general | 4 |
| Navigating statistical data | 6 | 5 | Can't find topic that inspires passion | 5 | 5 | Nothing to gain from doing research | 5 |
| Motivation | 5 | 6 | Scholarship teaching is focused a sister program; my program gets zoom, which doesn't work as well and feels like an afterthought | 6 | 6 | Difficult to find opportunities / no projects of interest available |  |
| Don't enjoy writing process of scholarly work | 7 | 7 | Lack of training: general | 7 | 7 | Lack of (personal) experience |  |
| Recruitment | 11 | 8 | Lack of knowledge: statistics | 8 | 8 | Lack of guidance: general |  |
| Knowing how to utilize mentors | 8 | 9 | Hard to find others to work with on a project | 9 (tie) | 9 (tie) | Lack of guidance: process / timeline |  |
| Funding | 13 | 10 | Lack of direction | 9 (tie) | 9 (tie) | Lack of motivation |  |
| Not sure how to start | 10 | 11 | Lack of training: study design | 9 (tie) | 9 (tie) | Lack of support: IRB takes months to get approval |  |
| Inexperience | 9 | 12 | No mentors | 9<br>(tie) | 9<br>(tie) | Limited resources: can't access articles that require payment |  |
| Lack of expertise | 12 | 13 | Hard to divide labor | 13 | 13 | Limited resources: general |  |
| Lack of ongoing projects of interest | 14 | 14 | Lack of support | 14 | 14 | Limited resources: lack of ancillary support |  |
|  |  |  | Lack of knowledge: general | 15 | 15 | Limited resources: lack of mentors |  |
|  |  |  |  |  |  | Limited resources: lack of statistician support |  |
|  |  |  |  |  |  | Limited supervision and coaching |  |
|  |  |  |  |  |  | Poor preceptor experience / conflict with faculty |  |
|  |  |  |  |  |  | Research is not a priority compared to other resident duties |  |
|  |  |  |  |  |  | Time: balance with work/life |  |
|  |  |  |  |  |  | Time: decreased residents to cover call |  |
|  |  |  |  |  |  | Time: general (not enough time) |  |
|  |  |  |  |  |  | Time: schedule changes |  |
|  |  |  |  |  |  | Unclear instructions from institution |  |

**Table 3.** Rank Biased Overlap (RBO) comparison between ranked barriers at different locations and within different specialties. Higher RBO correlates with more similarity in top-ranked items between two lists. RBO was higher within health systems than within specialties.

| Weighting Parameter | Top 5 Ranks' Weights | RBO |  |  |  |  |
| --- | --- | --- | --- | --- | --- | --- |
|  |  | S2-A and S2-B Family Medicine | S2-A Surgical and Non-Surgical | S3 Surgical and Non-Surgical | S2-A and S3 State Surgical | S2-A and S3 Non-Surgical |
| 0.9 | 67.2% | 0.4998 | 0.4138 | 0.4778 | 0.4073 | 0.3867 |
| 0.8 | 86.09% | 0.5540 | 0.5368 | 0.5615 | 0.5017 | 0.4687 |
| 0.7 | 94.12% | 0.6110 | 0.6298 | 0.6382 | 0.5833 | 0.5482 |
| 0.6 | 97.67% | 0.6681 | 0.7023 | 0.7047 | 0.6550 | 0.6251 |
| 0.5 | 99.18% | 0.7252 | 0.7614 | 0.7619 | 0.7199 | 0.6987 |
| 0.4 | 99.76% | 0.7822 | 0.8125 | 0.8126 | 0.7805 | 0.7680 |

### System 2-A and System 3 surgical and non-surgical residency programs

950 residents were sent the survey between the two sites with 98 completing the first survey (10% response rate) with 44 completing both subsequent rounds (55% dropout rate). Table 4 shows the barriers of S2-A Surgical, S2-A Non-Surgical, S3 Surgical, and S3 Non-Surgical. The top ranked barrier for all was lack of time. Table 3 shows the RBO for weighting parameters from 0.4 to 0.9 comparing these groups along with the family medicine residency comparison.

**Table 4.**
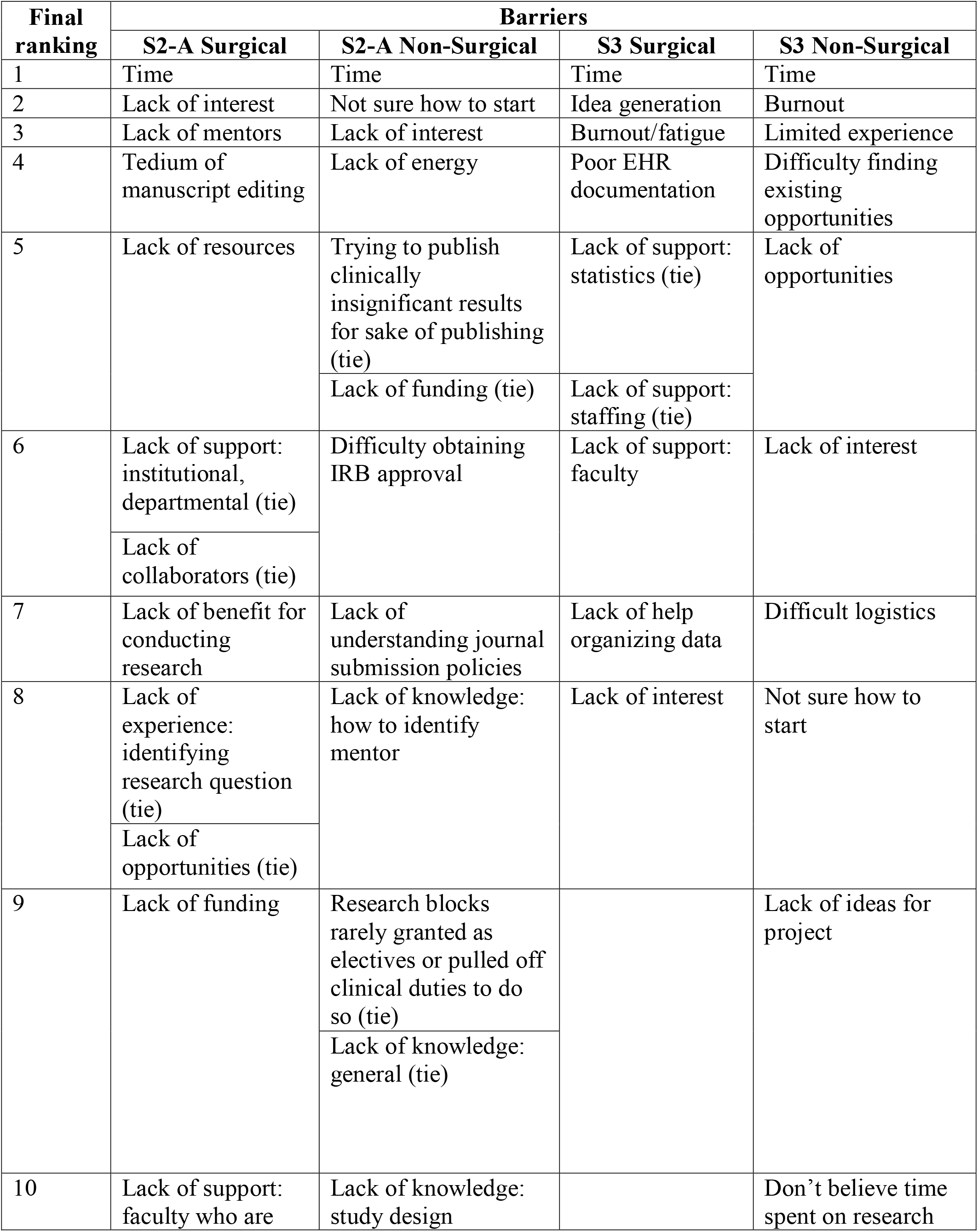
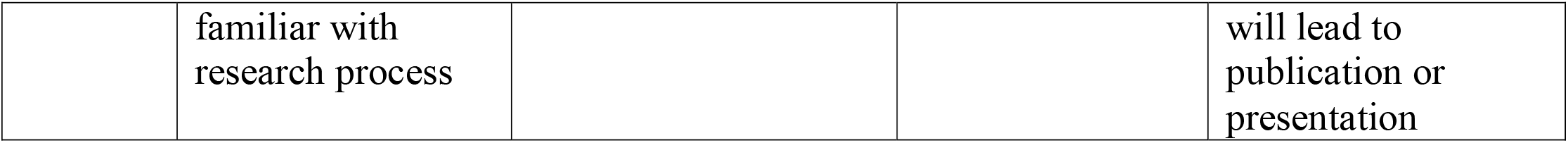
Residents’ ranked barriers to scholarly activity per site and specialty category using Delphi method.

## Discussion

To our knowledge, this is the first use of the Delphi and RBO techniques to evaluate barriers to scholarly activity among residents, fellows, and/or medical faculty. We demonstrate that the Delphi method allows programs to generate ranked lists of perceived scholarly activity barriers through the S1 study. While generation of such a list is useful in and of itself, comparison of lists between programs and specialties can allow administrators to glean best-practices for addressing barriers. If barriers are similar among the same specialty but differ by location, then specialty-specific organizations could play a role in addressing barriers. Conversely, if barriers are similar by location but different by specialty, then site- or system-specific interventions could be implemented.

To maintain fidelity, we limited our consolidation, keeping “Time,” “Prioritizing other learning experiences” and “Energy during residency when exhausted and busy” as distinct barriers. However, all of those may be considered “time” issues. Time was the top barrier for all resident-only groups (all S2 and S3 programs surveyed) (Tables 2 and 4).

Interestingly, the mixed cohort at S1 ranked time third (Table 1). This may be a reflection of both the persistence of time as a barrier through advanced fellowship and faculty and the risk of dilution in Delphi studies that include widely diverse populations. Given that all other resident groups identified time as their top barrier, it is reasonable to posit that S1 residents also did this, but their ranking may have been offset by non-residents, who might have more time built into their work schedules to engage in scholarship. Institution-wide Delphi studies to identify perceived barriers may prove less useful than studies that include a narrower range of participants.

Time was identified as the top resident barrier in a recent study, in which lack of funding was also identified as the second highest need. ^11^Residents in this study did not see lack of funding as a significant barrier. S2-B and S2-C Family Medicine, S3 Surgical, and S3 Non-Surgical residents did not even identify funding as a barrier (Tables 2, 4). S2-A Family Medicine residents ranked funding 11^th^ out of 14 (Table 2); S2 Surgical residents ranked funding 9^th^ out of 10; S2 Non-Surgical residents ranked funding as 5^th^ out of 10 (Table 4). This highlights the value of ranking in identifying barriers to help programs focus limited resources. It is not surprising that lack of funding was not identified as a major barrier in this study, since most of our participants were trainees, and would typically not be the ones seeking grant funding.

RBO enabled comparisons between groups with different barriers, and through its weighting parameter enabled the focus of comparison on top barriers. This is particularly useful for large organizations with many residencies, as it can aid in identifying the top barriers that are common across residency types.

RBO is a similarity measure that uses weights for each rank position. It judges the similarity of ranking lists without exact matches, and it allows for assigning greater weight to top ranks. In the context of RBO, the “p” is a tunable parameter (with values between 0 and 1), that can be used to determine the contribution of the top ranks to the final value of the RBO similarity measure. The value of the “p’ parameter is chosen based on the amount of contribution desired from the top results. The resulting RBO measure is also a number between 0 and 1, where 0 indicates that the lists are disjointed and 1 indicates perfect overlap. Though there are no defined cut-offs, an RBO of 0.8 is would be considered high overlap, indicating similar rankings. ^10^

## Limitations

A challenge of using the Delphi methodology for each resident group (or even year group) separately is the decrease in sample size due to the significant drop-out rate. For our data, we found the mixed group at S1 too broad. This could explain the observation that “time” as a perceived barrier ended up with a lower ranking.

Individual residencies had very high dropout rates, the S2-C family medicine program to such an extent that a third Delphi round was not completed. Our comparisons of surgical versus non-surgical residents at two sites appeared to be a reasonable approach to address both extremes. Residents identified time as the time barrier across all four groups, consistent with the literature, but the top five barriers varied by institution and residency type.

Our data shows high RBO across all p parameter values for the two Family Medicine programs (Table 3). The top five barriers for S2-A Surgical versus S2-A Non-Surgical, and S3 Surgical versus S3 Non-Surgical, indicate a high overall overlap (0.81 for both, Table 3). However, while we might expect surgical resident barriers to be similar to other surgical resident barriers, we do not see that to the same degree as overall resident similarity. S2-A Surgical versus S3 Surgical has an RBO of 0.78 – high, but not as high as the intra-institution scores. Similarly, S2-A Non-Surgical versus S3 Non-Surgical has an RBO of 0.77 (Table 3). We interpret this to mean that while there is overall high similarity among residents’ perceived barriers to scholarly activity, these barriers vary more by institution than specialty. Hence, solutions may better be sought which are institution-specific versus specialty-specific.

Our study highlights the challenges of resident retention in studies over time and may point towards a general lack of prioritization of scholarly activities. Although Family Medicine residents had dedicated time and financial incentives, only 10 out of 61 residents completed the surveys. Future studies with larger or more residency programs and better follow-up with previously sent surveys may address this issue.

Medical center size appears to have minimal impact on perceived barriers to scholarship, with high overlap between S1 (204-bed facility), S2-A (628-bed facility), S2-B (260-bed facility), and S3 (663-bed facility). Many barriers may be addressed through administrative interventions or education. The educational gaps that would be easiest to address are those requiring only information about how to access existing support. Other gaps may be addressed with focused education modules housed through a central organization, like the Society of Teachers of Family Medicine resource library.^12^

Study limitations include a low response rate and high drop-out rate. Several of the residencies (the three Family Medicine programs, S2-A Surgical and S2-A Non-Surgical programs) are within the same system, which may limit generalizability. However, this limitation is mitigated by data from two other health systems and the fact that two of the Family Medicine programs are physically remote from S2-A, with the bulk of their training at different hospitals. Also, our study was completed during the COVID-19 pandemic, which may have had an impact on participants’ perceptions and their responses.

## Conclusion

Major barriers to scholarly activity among physicians include lack of time and other challenges, such as understanding the IRB process and inadequate mentorship. Several identified barriers can be addressed with focused educational interventions. Using the Delphi method, we were able to identify ranked barriers to scholarly activity among medical learners and compare them using the RBO technique. Our study results can help guide educational efforts at participating institutions, but further research is needed to assess the generalizability of these preliminary findings.

## Data Availability

All data produced in the present study are available upon reasonable request to the authors

